# Deep brain stimulation in the bed nucleus of the stria terminalis: a symptom provocation study in patients with obsessive-compulsive disorder

**DOI:** 10.1101/2021.03.12.21253450

**Authors:** Kelly Luyck, Chris Bervoets, Choi Deblieck, Bart Nuttin, Laura Luyten

## Abstract

**Background:** Deep brain stimulation (DBS) is an emerging therapy for treatment-resistant obsessive-compulsive disorder (OCD), and several targets for electrode implantation and contact selection have been proposed, including the bed nucleus of the stria terminalis (BST). Selecting the active electrode contacts (patients typically have four to choose from in each hemisphere), and thus the main locus of stimulation, can be a taxing process. Here, we investigated whether contact selection based purely on their neuroanatomical position in the BST is a worthwhile approach. For the first time, we also compared the effects of uni- versus bilateral BST stimulation.

**Methods:** Nine OCD patients currently receiving DBS participated in a double-blind, randomized symptom provocation study to compare no versus BST stimulation. Primary outcomes were anxiety and mood ratings in response to disorder-relevant trigger images, as well as ratings of obsessions, compulsions, tendency to avoid and overall wellbeing. Furthermore, we asked whether patients preferred the electrode contacts in the BST over their regular stimulation contacts as a new treatment setting after the end of the task.

**Results:** We found no statistically significant group differences between the four conditions (no, left, right and bilateral BST stimulation). Exploratory analyses, as well as follow-up data, did indicate that (bilateral) bipolar stimulation in the BST was beneficial for some patients, particularly for those who had achieved unsatisfactory effects through the typical contact selection procedure.

**Conclusions:** Despite its limitations, this study suggests that selection of stimulation contacts in the BST is a viable option for DBS in treatment-resistant OCD patients.

## Introduction

Obsessive-compulsive disorder (OCD) is a burdensome psychiatric disorder, characterized by persistent, often anxiety-provoking obsessions and time-consuming compulsions (1). It affects 2% of the population, and although pharmacological or cognitive behavioral therapy can reduce symptoms for many patients, about 10% remains severely incapacitated (2, 3). Luckily, treatment opportunities are emerging also for these treatment-resistant patients (4). One such ‘last-resort’ option is deep brain stimulation (DBS). First proposed about 20 years ago (5), this approach has been adopted worldwide, with encouraging clinical results (6-10). Nevertheless, the treatment is still new in terms of the number of patients that have been treated (a few hundred in total). Thus, it is of paramount importance that we scrutinize and attempt to refine this emerging treatment option, both for our current patients and for future ones. One aspect deserving consideration is the locus of stimulation, e.g., through the choice of active contacts from the 4 available contacts in each hemisphere, or even through an adjusted site of implantation for new patients. At present, various neural targets are being used for DBS in OCD patients, e.g., the anterior limb of the internal capsule, nucleus accumbens and the subthalamic nucleus (7-9). Very recently, an elegant connectome analysis identified a DBS site (white matter tract) that may result in optimal clinical improvement, but which still awaits further validation in clinical trials (11). Here, we used an alternative approach rooted in current knowledge of the neuroanatomy of (pathological) anxiety. Specifically, growing evidence suggests that the bed nucleus of the stria terminalis (BST) could be a promising target region (12-15). This is supported by a comparison of the therapeutic effects of DBS in the BST versus internal capsule in a prior study (16). Also in a more recent clinical trial, the active contacts were located in or adjacent to the BST in several patients (17). Furthermore, preclinical research has provided compelling evidence for a role of the BST in anxiety (18, 19), and there are even indications that high-frequency electrical stimulation in the BST can attenuate anxiety and compulsive behavior in rodent models (20, 21). Taken together, the BST is clearly a region of interest for DBS in OCD. Interestingly, there are some indications that BST functioning may be somewhat asymmetric (22, 23). We found, for instance, that the left BST is more strongly activated than its right counterpart in a rat model of anxiety (24). In addition, visual inspection of published imaging data suggests that BST activity mostly originated from the left hemisphere in human studies on anxiety and threat monitoring (25, 26). Finally, we occasionally have patients that fare better with unilateral stimulation (16). Thus, although DBS is usually applied bilaterally, it seems worthwhile to examine the effects of unilateral stimulation.

In this study, we investigated the effects of unilateral and bilateral DBS in the BST in a double-blind, randomized design. To minimize discomfort for our burdened patient population, we designed a computerized symptom provocation study with 4 stimulation conditions (left, right, bilateral BST or no stimulation) with a predetermined stimulation frequency and pulse width. Patients were asked to rate the effects under the 4 conditions on anxiety/stress, depressive feelings, obsessions, compulsions, avoidant tendency and overall wellbeing. We focused not only on OCD core symptoms obsessions and compulsions, but first and foremost on anxiety and mood, as they tend to fluctuate more rapidly when stimulation settings are changed (7, 16, 27). Apart from providing fundamental insights in the effects of unilateral versus bilateral BST stimulation for the first time, this study is also a first endeavor to select stimulation contacts purely guided by their anatomical position and a brief, standardized symptom provocation task. Finding optimal stimulation parameters (i.e., choice of contacts, voltage, frequency and pulse width) is often a taxing and time-consuming series of trials and errors in the psychiatrist’s office, and it can take weeks to months to find satisfactory settings (8, 28-30). In the current study, we try out a fundamentally different approach, based on the neuroanatomical location of the contacts, which has the potential to significantly streamline this process.

## Materials & Methods

This study was approved by the Ethics Committee of the University Hospitals Leuven (S62175) and preregistered on clinicaltrials.gov (NCT03894397). All data are available on Figshare via the Open Science Framework: https://dx.doi.org/10.17605/osf.io/enwju. Additional information is available in the ***Supplement***.

### Patient selection

Patients were recruited from the cohort of 41 OCD patients who were previously implanted with a DBS system at University Hospitals Leuven and had received at least 3 months of electrical stimulation. We excluded patients who were currently not being stimulated and patients who only display symptoms in specific environments (e.g., in their own home), as we would not be able to evoke symptoms in a hospital setting. Patients who did not speak Dutch (and could therefore not understand the questionnaires and audio fragments) or (elderly) patients who suffered from cognitive impairment were also excluded. In the remaining patient pool, we evaluated electrode location by merging previously acquired pre-op MRI and post-op CT images (iPlan Net, Brainlab, Munich, Germany) for every patient (16). For both hemispheres, we determined the electrode contact that was optimally located in (or maximally 4 mm from the anatomical outline of) the BST. Patients with bilateral electrode contacts in the BST (or within 4 mm) were considered eligible for study participation. Sixteen patients were eligible and were informed about the planned study and invited for participation. Thirteen patients initially provided their informed consent, nine of which (6 women, 3 men, 30-60 years old) eventually took part in the study. Patients 1 and 9 were previously included in (16), and patients 3, 4, 6 and 8 in (17). In a prior study with a long-duration (3 months) double-blind crossover phase comparing stimulation versus no stimulation, differences in anxiety and depression symptoms (measured with the Hamilton Anxiety and Depression Rating Scale) reached Cohen’s *d*_*z*_ effect sizes of 1.7-1.8 (16). Assuming these effect sizes, paired t-tests with 9 subjects would provide a power of >99% to detect differences between two stimulation conditions. Differences with an effect size of at least 1.1 should be detected with a power of >80%.

### Procedure

Baseline measurements were acquired while the patient was receiving DBS as usual, maximally 10 weeks before the symptom provocation task. Patients were asked to fill out self-report scales of the BDI (Beck Depression Inventory) and STAI (State-Trait Anxiety Inventory). In addition, Y-BOCS (Yale-Brown Obsessive Compulsive Scale, to assess symptom severity) and laterality questionnaires were administered by the psychiatrist. See ***Fig. 1*** for an overview of the study protocol.

**Fig. 1:**
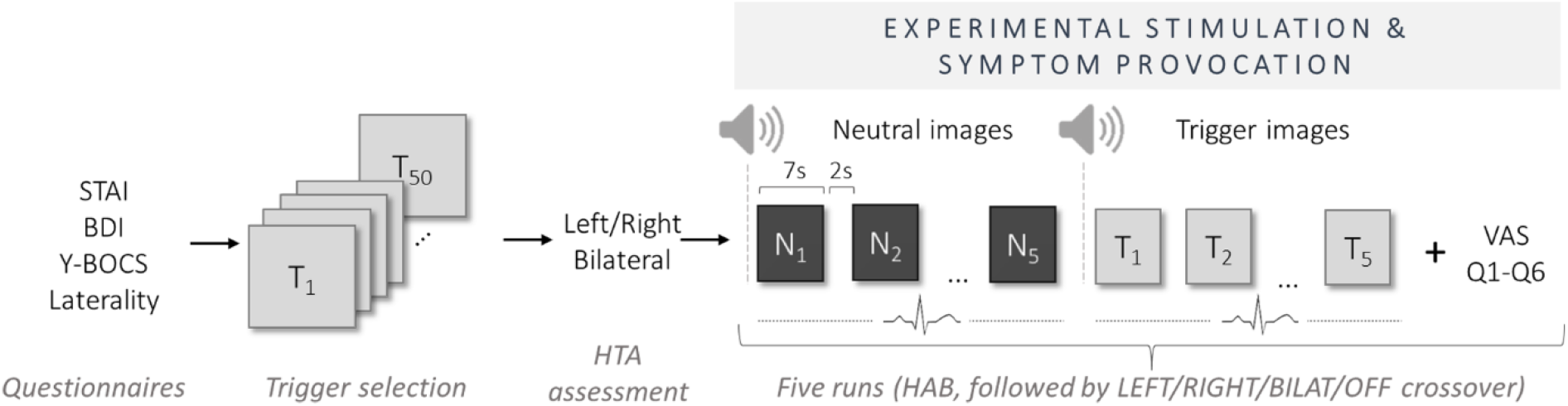
Overview of the study protocol. BDI: Beck Depression Inventory; BILAT: bilateral; HAB: habituation; HTA: highest tolerable amplitude; Q1-Q6: question 1-6; STAI: State-Trait Anxiety Inventory; VAS: visual analogue scale; Y-BOCS: Yale-Brown Obsessive Compulsive Scale.

On the day of the symptom provocation task, patients were presented with a series of 50 images from the Maudsley Obsessive-Compulsive Stimuli Set (MOCCS) that were relevant to their OCD subtype (i.e., washing or checking obsessions and compulsions) in the psychiatrist’s office (31). Each image was shown for 7 seconds and patients were asked to quantify the level of anxiety/stress provoked by each individual picture using a visual analogue scale (VAS) on the computer screen. The VAS was a straight horizontal line without numerical ratings nor gradations, with the ends defined as the extreme limits of the parameter under investigation (e.g., ‘not at all’ to ‘very much’), which was recorded by the computer as a value from 0 to 100%. The five images that evoked the strongest level of anxiety, hereafter referred to as trigger images, were selected for the crossover phase of the study. All images, accompanying audio instructions and rating scales were presented using Affect 5.0 software (32).

Next, in the laboratory, patients were seated in front of a computer, electrocardiography (ECG) electrodes were attached, and cameras were positioned for observation of the patient and psychiatrist. We then determined the highest tolerable amplitude (HTA), i.e., the maximal amplitude that was tolerated by the patient without generating any persistent side effects. The stimulation amplitude was gradually increased until non-transient side effects (e.g., tingling sensations at various locations, flushing, transpiration, etc.) were observed or reported by the patient. At this point, the amplitude was lowered until side effects ceased. This procedure was performed for stimulation of the left and right (random order), and finally bilateral BST.

In the following phase, the effects of stimulation were assessed in a computerized symptom provocation task. Patients were first evaluated while being stimulated with their regular DBS settings during the Habituation phase (HAB), which may or may not comprise electrode contacts in the BST ***(Supplement, Table S3)***. The aim was to familiarize patients with the computer task before applying the experimental stimulation parameters, and to quantify their effect during symptom provocation. The patient and psychiatrist were not blinded during the HAB phase. The patients were first presented with five neutral images that were completely unrelated to their obsessions, followed by the five trigger images that were determined previously. Both series of neutral and trigger images were introduced by an audio fragment that stated the nature of the images that would follow, i.e., first neutral and then trigger images. Following presentation of the five triggers, the patients filled out a VAS for the following symptoms: Q1: anxiety/stress; Q2: depressive feelings; Q3: urge to perform compulsions; Q4: obsessive thoughts; Q5: urge to avoid the situation; Q6: wellbeing ***(Supplement, Table S5)***.

Next, four experimental stimulation conditions were evaluated in a randomized order using the previously determined HTAs: left BST stimulation (LEFT), right BST stimulation (RIGHT), bilateral BST stimulation (BILAT) or no stimulation (OFF). During this part of the study, both the psychiatrist and the patient were blinded to the stimulation condition. The symptom provocation procedure with five neutral and five trigger images and VAS ratings was repeated for all four stimulation conditions, and a brief wash-out period of about 5 minutes was installed between every condition.

### Electrical stimulation

Patients included in this study were implanted (0.3 to 10.6 years ago, 6.8 on average) with bilateral 3391-28 or 3387-28 type leads, connected to an Activa PC or Activa RC system (Medtronic Inc, Minneapolis, MN, USA). Preoperative levels of OCD, anxiety and depressive symptoms are shown in ***Table S1 (Supplement)***.

Throughout the current study, adjustment of parameters was done by an unblinded researcher, using the Medtronic Clinician Programmer tablet. During HAB, patients were stimulated with their regular contact configuration, frequency, pulse width and amplitude ***(Supplement, Table S3)***. For experimental BST stimulation, the contact that was closest to the center of the BST (or within 4 mm of the anatomical BST outline) was used as a cathode, with the anode being the adjacent contact dorsal to it. Pulse width and frequency were fixed at 450 µs and 130 Hz, respectively, during all experimental conditions and for each patient. The pulse width was selected based upon our clinical experience in parameter optimization, which suggests that relatively long pulse widths tend to produce better therapeutic effects in OCD patients. As described above, amplitudes were set as high as possible, without inducing side effects ***(Supplement, Table S4)*** (33). Note that the safety threshold of 30 µC/cm^2^ was never exceeded.

### Statistical analyses

Statistical analyses were performed and graphs were created using GraphPad Prism (v7.02). Data are shown as individual scores and/or means with SEM. Responses to the questions on the VAS scales were compared with paired t-tests for the HAB versus OFF condition and with one-way repeated-measures ANOVAs for the comparison of OFF, BILAT, LEFT and RIGHT stimulation conditions. For exploratory analyses, patients were divided into two subgroups according to their baseline Y-BOCS. Responses on the VAS scales were then analyzed with two-way repeated-measures ANOVA for the comparison of HAB, OFF and BILAT conditions, and followed up with Tukey’s posthoc tests.

## Results

### Patient and stimulation characteristics

Baseline measurements indicated a rather large variability in the severity of OCD (Y-BOCS ranging from 5 to 28), anxiety (STAI Trait/State ranging from 27 to 79) and depressive symptoms (BDI ranging from 0 to 50) ***(Supplement, Table S2 and Fig. S1)***.

The BST stimulation contacts (***Fig. 2***) were located in the central, lateral or posterior division of the BST for 8 patients, and for Pt06, contacts were within the predefined range of 4 mm, with the cathode centers positioned in the lateral hypothalamic area. For all patients, except Pt01, BST stimulation was done with the most ventral contact C0 (C1 for Pt01). Notably, for all patients, there was a change in contacts that were used for BST stimulation as compared with their regular stimulation settings (HAB) ***(Supplement, Table S3)***. For one patient (Pt07), the cathodes were identical in both cases, but the anodes, frequency, pulse width and voltages differed, implying that the electrical field would be different in both conditions. For their regular stimulation, only 3 out of 9 patients received bipolar stimulation, whereas for BST stimulation in the present study, all received bipolar stimulation with the adjacent contact as anode. Highest tolerable stimulation amplitudes in left, right and bilateral BST did not differ significantly ***(Supplement, Fig. S2)***.

**Fig. 2:**
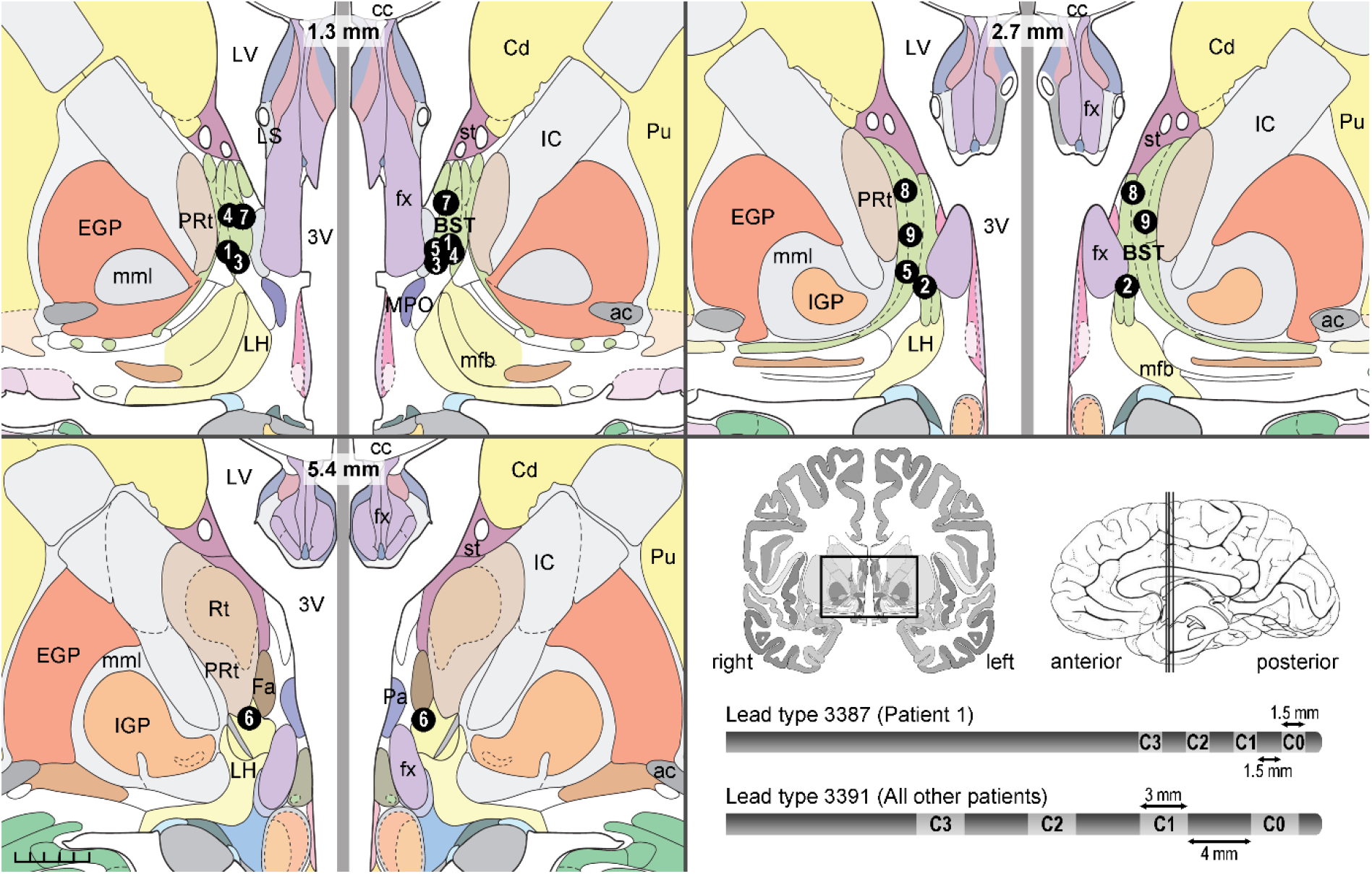
Electrode contact locations for stimulation of the BST. Coronal brain slices showing the location of the center of cathodes selected for stimulation of the BST. Contacts are depicted as black circles and include the patient number. On top of each of the three coronal slices, the position posterior to the anterior commissure is specified, and right and left hemispheres are depicted adjacently, in accordance with the radiological convention (right hemisphere shown on the left side and vice versa). In the bottom left corner is a 5-mm scale. Instead of displaying complete coronal slices, a detailed window is shown, dorsally bordered by the corpus callosum and ventrally extending 10 mm below the intercommissural plane, as indicated in the bottom right panel. A sagittal view of the brain with indication of the relevant coronal slices is also displayed. Additionally, the lead types used in this study are shown, with indication of the contact numbering (C0-C3). ac: anterior commissure; BST: bed nucleus of the stria terminalis; cc: corpus callosum; Cd: caudate nucleus; EGP: external globus pallidus; Fa: fasciculosus nucleus; fx: fornix; IC: internal capsule; IGP: internal globus pallidus; LH: lateral hypothalamic area; LS: lateral septal nucleus; LV: lateral ventricle; mfb: medial forebrain bundle; mml: medial medullary lamina of globus pallidus; MPO: medial preoptic nucleus; Pa: paraventricular hypothalamic nucleus; PRt: prereticular zone; Pu: putamen; Rt: reticular thalamic nucleus; st: stria terminalis; 3V: third ventricle. Images are adapted from Mai’s Atlas of the Human Brain (34).

### Quantitative analysis of the symptom provocation task

Responses to the questions probing feelings of anxiety/stress, depression, obsessions, compulsions, avoidance and wellbeing during the different stimulation conditions are shown in ***Fig. 3*** (individual patient profiles can be found in ***Supplement, Fig. S3***). On average, the open-label HAB condition had numerically better ratings than the double-blind OFF condition on all 6 responses, but these differences were not statistically significant (***Table 1***). Furthermore, one-way repeated-measures ANOVAs with Greenhouse-Geisser correction showed no significant differences across the 4 conditions (***Table 1***). There was no indication that unilateral stimulation outperformed bilateral, or even no stimulation. Visual inspection of the data (***Fig. 3***) does suggest that, on average, bilateral BST stimulation (BILAT) had numerically better effects than no stimulation (OFF), and was comparable to open-label stimulation with the patient’s regular stimulation settings (HAB). These effects were, however, not statistically supported in this sample of only 9 patients.

**Table 1:** Statistical results for responses on visual analogue scales. Paired t-tests (comparison of HAB and OFF) and one-way repeated measures ANOVAs (comparison of double-blind crossover conditions OFF, BILAT, LEFT and RIGHT).

|  | Comparison HAB and OFF | Comparison OFF, BILAT, LEFT and RIGHT |
| --- | --- | --- |
| **Q1 – Anxiety/stress** | $t(8) = 0.43, p = .68$ | $F(2.36, 18.87) = 1.33, p = .29$ |
| **Q2 – Depression** | $t(8) = 1.30, p = .23$ | $F(2.26, 18.05) = 0.48, p = .65$ |
| **Q3 – Obsessions** | $t(8) = 0.99, p = .35$ | $F(2.78, 22.20) = 1.05, p = .39$ |
| **Q4 – Compulsions** | $t(8) = 1.07, p = .31$ | $F(2.89, 23.13) = 0.63, p = .60$ |
| **Q5 – Avoidance** | $t(8) = 1.46, p = .18$ | $F(2.13, 17.07) = 0.76, p = .49$ |
| **Q6 – Wellbeing** | $t(8) = 0.29, p = .78$ | $F(2.01, 16.09) = 0.26, p = .78$ |

**Fig. 3:**
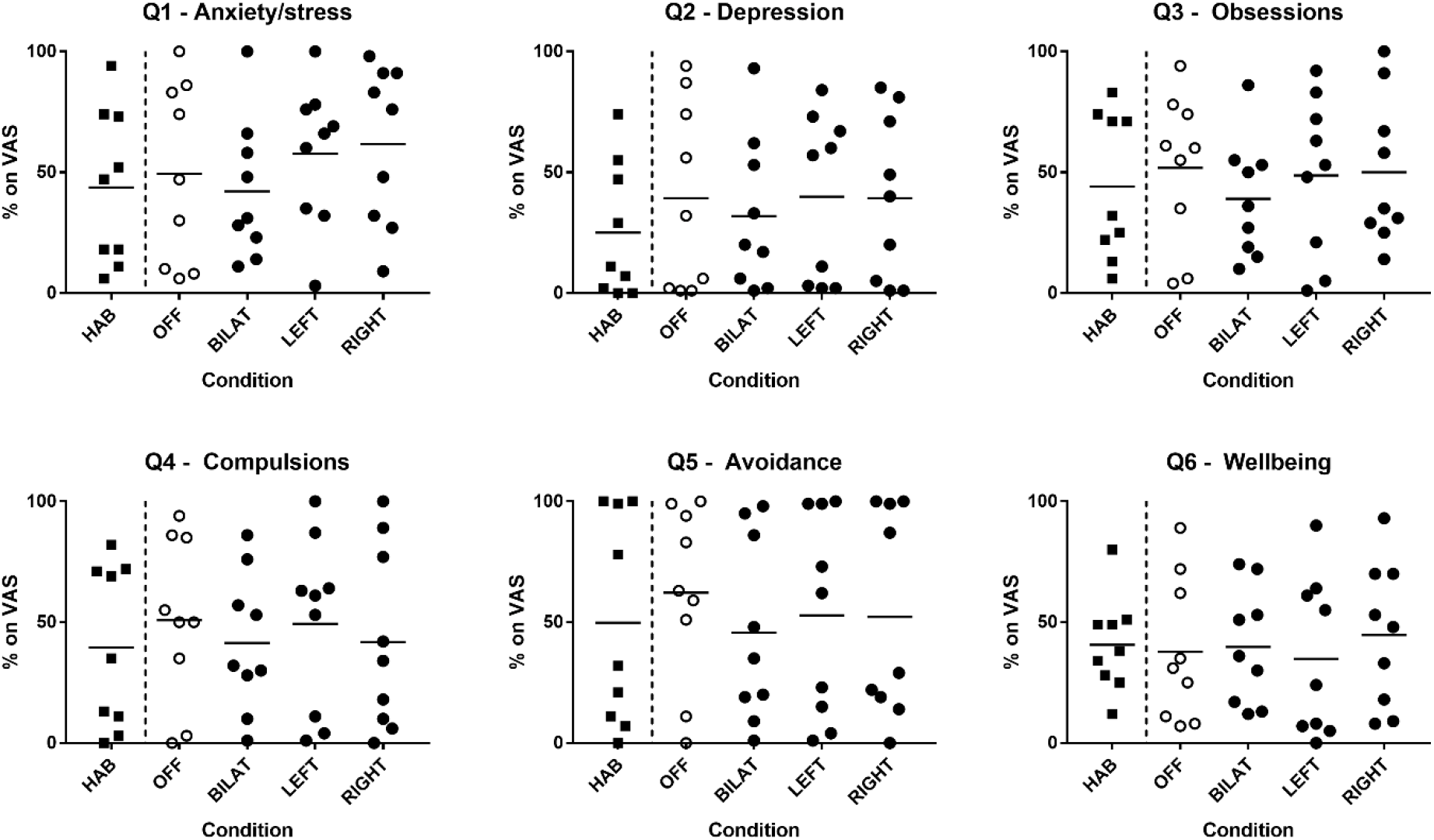
Individual data and means of 9 patients are shown for the responses to six questions related to feelings of anxiety/stress (Q1), depression (Q2), obsessions (Q3), compulsions (Q4), tendency to avoid (Q5) and feeling of wellbeing (Q6). Data from the Habituation (HAB) phase are separated from the other measurements with a vertical dashed line because this open-label phase was not part of the crossover. During the crossover, patients went through 4 double-blind, randomized phases (OFF (no stimulation), BILAT (bilateral), LEFT and RIGHT stimulation). Symbols for the OFF condition are shown in white since it is the only condition without any electrical brain stimulation. VAS: visual analogue scale.

As illustrated in ***Fig. 3***, there was a large variability in the responses of the patients. One reason for this variability may be baseline differences in how severely patients were affected by their disorder, and the accompanying room for improvement. Although all patients had lower Y-BOCS scores compared with the preoperative level (52% improvement on average), the individual numbers ranged from 20% to 86%. For exploratory purposes, we therefore conducted analyses in two subgroups (***Fig. 4***); those with mild OCD at the start of this study (Y-BOCS up to 15, n = 4) and those with moderate to severe OCD (Y-BOCS: 16-40, n = 5) (35, 36).

**Fig. 4:**
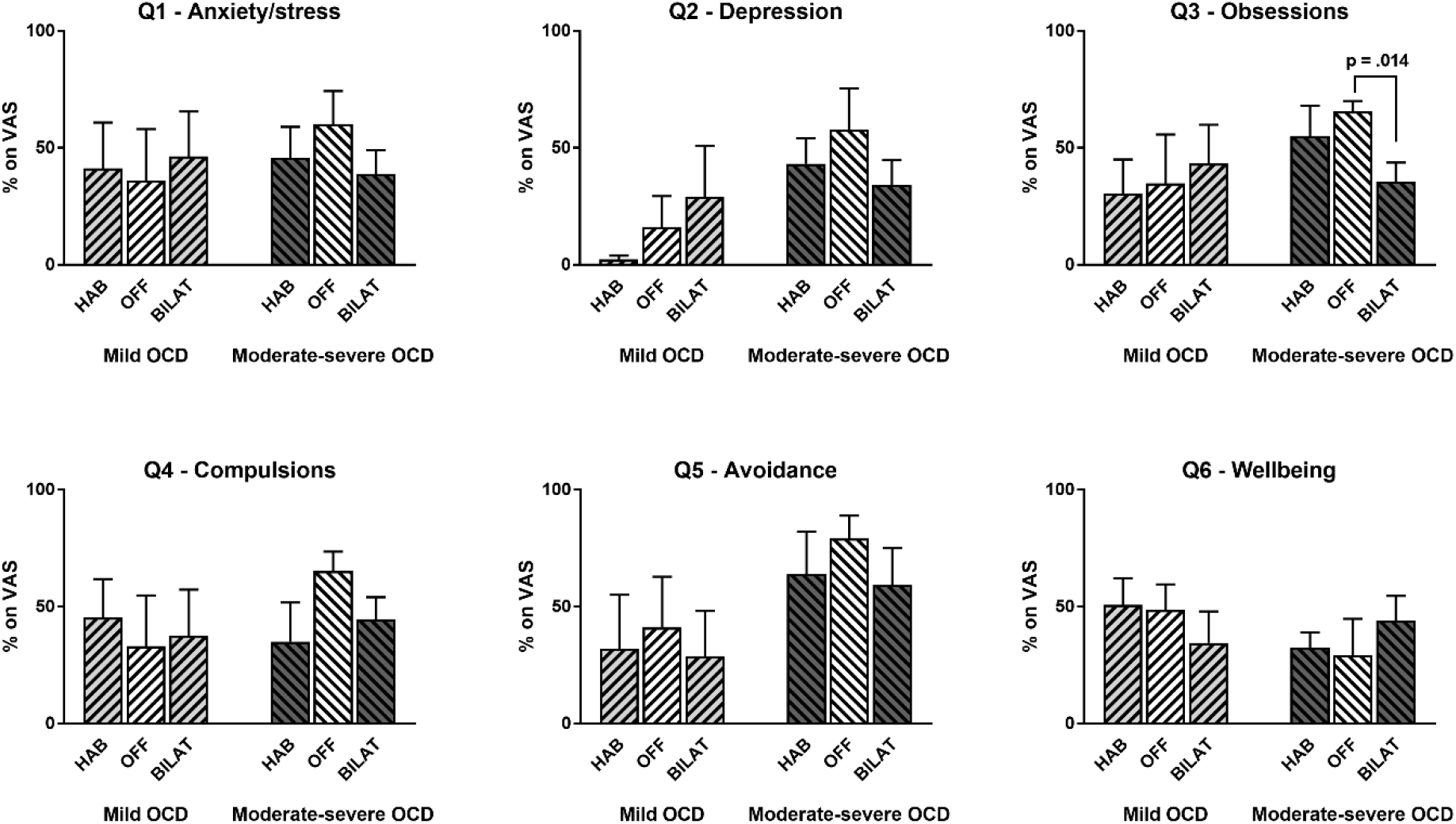
Responses (mean and SEM) to six questions related to feelings of anxiety/stress (Q1), depression (Q2), obsessions (Q3), compulsions (Q4), tendency to avoid (Q5) and feeling of wellbeing (Q6). Data are shown for two subgroups, one with mild OCD at baseline (n = 4) and one with moderate-severe OCD (n = 5). Responses are compared for stimulation as usual (HAB), no stimulation (OFF) and bilateral BST stimulation (BILAT). VAS: visual analogue scale.

We compared no stimulation (OFF) with stimulation as usual (HAB) and bilateral stimulation of the BST (BILAT) using a two-way repeated-measures ANOVA, followed up with Tukey’s posthoc comparisons of the 3 conditions within each subgroup (***Table 2***). Follow-up tests for the responses to Q3 regarding obsessions in the subgroup with moderate-severe OCD indicated a significant improvement of obsessions with bilateral BST stimulation compared to no stimulation (mean improvement of 30%, 95%-confidence interval [6.2%; 53.8%], p = .014).

**Table 2:** Statistical results for responses on visual analogue scales in subgroups with mild versus moderate-severe OCD at baseline. Two-way repeated-measures ANOVAs comparing HAB, OFF and BILAT. One interaction, for the question regarding obsessions, was statistically significant and is indicated in bold.

|  | Main effect of subgroup | Main effect of stimulation condition | Interaction of subgroup and stimulation condition |
| --- | --- | --- | --- |
| **Q1 – Anxiety/stress** | $F(1,7) = 0.13, p = .73$ | $F(2,14) = 0.14, p = .87$ | $F(2,14) = 1.06, p = .37$ |
| **Q2 – Depression** | $F(1,7) = 3.44, p = .11$ | $F(2,14) = 0.91, p = .42$ | $F(2,14) = 1.94, p = .18$ |
| **Q3 – Obsessions** | $F(1,7) = 0.88, p = .38$ | $F(2,14) = 1.30, p = .30$ | **$F(2,14) = 4.57, p = .03$** |
| **Q4 – Compulsions** | $F(1,7) = 0.26, p = .63$ | $F(2,14) = 0.55, p = .59$ | $F(2,14) = 2.56, p = .11$ |
| **Q5 – Avoidance** | $F(1,7) = 1.98, p = .20$ | $F(2,14) = 2.44, p = .12$ | $F(2,14) = 0.15, p = .87$ |
| **Q6 – Wellbeing** | $F(1,7) = 0.61, p = .46$ | $F(2,14) = 0.04, p = .96$ | $F(2,14) = 1.28, p = .31$ |

Visual inspection of the graphs (***Fig. 4***) suggests that the pattern that was observed in the full dataset – i.e., comparable ratings with regular (HAB) and with bilateral BST stimulation, with both conditions being numerically (but not significantly, except for the question related to obsessions) better than the OFF condition – is most evident in patients with moderate-severe OCD complaints at baseline.

Laterality measures indicated that 3 patients were left-handed and 6 were right-handed. Exploratory analyses found no evidence for an interaction between handedness and the effects of unilateral BST stimulation on the responses to the six questions ***(Supplement, Fig. S4)***.

For exploratory purposes, ECG measurements were collected during image presentation throughout the crossover. No significant differences were found between the conditions ***(Supplement, Fig. S5-6)***.

### Qualitative assessment of the symptom provocation task

An elaborate qualitative analysis will be published elsewhere. Here, we include a brief overview of the observations of one experimenter (KL) and side effects that were mentioned by the patients themselves during the task (while blinded to the stimulation condition), and which may be relevant for interpretation of the quantitative data.

Several patients (n = 5) were (very) tense about the prospect of participating in the experiment, primarily because they would have to complete a computer task and were not in control of their stimulation settings. The most common side effects of uni- or bilateral BST stimulation were feeling (unpleasantly) warm (n = 7), (excessive) sweating (n = 4) and a strange or absent feeling (n = 3). A few patients presented with nausea (n = 2), heart palpitations (n = 2) or sporadic urge to move (n = 2). Although often transient, and attenuated by lower stimulation voltages, it is noteworthy that these side effects were perceived as surprising, and even worrisome, by some of the patients, especially those who had been on stable stimulation settings for years. This resulted in the OFF phase being experienced as less aversive (notwithstanding the presentation of trigger images). While uninformed about the stimulation condition, several patients (n = 5) clearly indicated that they were feeling calmer or better during the phase without stimulation.

### Switching to BST stimulation contacts after the study

Following the symptom provocation task, the majority of the patients (n = 6) decided to return to their regular stimulation contacts and monopolar stimulation, and were still using these settings one year later. Note that for all of them, the selected cathodes were adjacent to or even overlapping with the cathodes that were defined as being the closest to the center of the BST for the purpose of the current study. We can therefore assume that these patients received at least partial stimulation of the BST area, especially when considering a larger action radius of monopolar versus bipolar stimulation (16).

One patient (Pt08) switched to different settings after the study (but not to the contact settings that were used during the study) and was off stimulation one year later.

Two patients explicitly asked to change their active stimulation contacts to the ones that were used for bilateral BST stimulation during the task. Pt05 switched to bipolar stimulation with the cathode that was used during the study (C0), and still maintained these settings one year later. Pt07 was already being stimulated with the BST cathode (C0) before the start of the study, but now also decided to switch anodes (C3 to C1), so that the stimulation contacts were identical to those used during the task. This patient continued with these setting for 3 months, but then switched off the stimulator entirely for the next 9 months, due to external reasons unrelated to the stimulation effects. Notably, both patients were part of the moderate-severe subgroup (***Fig. 4***), which aligns with the assumption that neuroanatomically guided stimulation of the BST can be a worthwhile option to explore if there is still room for symptom improvement. Moreover, ***Fig. S3 (Supplement)*** suggests that both patients experienced beneficial effects when receiving bilateral versus no stimulation during the symptom provocation task.

## Discussion

We evaluated the effects of uni- and bilateral DBS in the BST in 9 OCD patients during an acute symptom provocation task, using a blinded crossover study design. The main hypothesis was that BST stimulation would improve ratings of anxiety/stress, mood and other symptoms compared with no stimulation. Although the group averages suggest an effect in this direction, particularly with bilateral stimulation, none of the group differences were statistically significant. For the subhypothesis that unilateral stimulation of the BST in the left hemisphere in particular would be beneficial, we found no evidence at all. Given the diversity of our small sample, we conducted a secondary analysis in which patients were divided in subgroups based on the severity of OCD at the start of the study, as measured by the Y-BOCS. This exploratory analysis suggested that bilateral BST stimulation may be beneficial particularly for patients with moderate to severe OCD, but only reaching a statistically significant improvement for the ratings pertaining to obsessions provoked by the image viewing task.

We should note that this trial has a few limitations which may explain the lack of statistical significance of the effects, and which may at the same time be worthwhile to take into account for future studies. First, it should be noted that the symptom provocation task may not have achieved its purpose in all patients. Although the triggers came from a validated set of images (31), several patients did not perceive the pictures as being very anxiety/symptom-provoking (as seen in the ratings of the image selection procedure and/or because they mentioned it during the actual task). In contrast, many did experience the computer task in itself to be quite stressful, thereby possibly overruling the potential of the images to trigger OCD symptoms. However, this overall high stress level among patients also suggests that using stronger (e.g., real-life) triggers may not be feasible in this population with (a history of) severe to extreme OCD (31, 37, 38).

Second, we should acknowledge that proper HTA determination was difficult in some patients, partially due to their general nervosity. Future studies should take into account that this can be a time-consuming process, particularly when patients are stimulated with very different settings than usual, including bipolar instead of monopolar stimulation, or long pulse widths (16, 29, 39). As a result, some side effects were still present during the task, and were perceived as rather aversive, unintentionally making the OFF phase a more pleasant condition. In the end, patients may have primarily rated their overall feelings and thoughts in each condition, rather than their responses to the trigger images.

Finally, it is worth mentioning that the standardized symptom provocation task was mainly included to improve the experimental design and scope of the study, but is probably not necessary for parameter optimization in daily clinical practice. Perhaps it could be useful, with more practice for the patients (so that the computer task itself feels less daunting) and/or with more aversive and varying trigger images.

Despite its limitations – which are largely inherent to this type of research – our study offers interesting take-home messages for clinicians who treat OCD patients with DBS.

Although no statistically significant differences were found at the group level, individual data inspection indicates that several patients experienced acute beneficial effects with BST stimulation. For instance, two thirds of the patients (n = 6) gave lower anxiety/stress and depression ratings (which were both preregistered as primary outcomes for this study) with uni- or bilateral BST stimulation than with their regular stimulation parameters. We should be careful when interpreting this comparison, as patients were unblinded during the regular stimulation condition, which was always tested first. Nevertheless, this does suggest that bilateral (n = 3), right-sided (n = 2) or left-sided (n = 1) BST stimulation may outperform stimulation settings that have been optimized for months/years. It also suggests that it may be worthwhile to consider unilateral or asymmetric (e.g., different voltages in both hemispheres) stimulation in OCD patients, something that is not often done (16, 29).

Furthermore, it is good to highlight that two patients who experienced such beneficial acute effects also requested to be stimulated at the contacts in the BST at study completion. This shows that DBS contact selection based purely on neuroanatomy is a viable method to aid in the challenging DBS optimization process. Here, this approach was tested after long-term stimulation and optimization with the standard “trial-and-error” procedure (8, 30), but we argue that it could be considered as one of the first steps of the selection process as well. Prior clinical and preclinical research have already shown that DBS targeted at the BST is a defensible choice (16, 20, 21, 40), and the present study provides new evidence to support this notion.

In conclusion, we found no statistically significant evidence in favor of uni- or bilateral BST versus no stimulation using a double-blind, randomized symptom provocation task. Additional analyses, as well as follow-up data, do however suggest that selection of the stimulation contacts purely based upon their location in the BST may be a worthwhile approach for some patients, particularly for those who have achieved unsatisfactory stimulation settings using the sometimes cumbersome, standard “trial-and-error” selection procedure.

## Data Availability

All data related to analyses presented in the manuscript are available on Figshare.

https://dx.doi.org/10.17605/osf.io/enwju

## Author contributions

Kelly Luyck and Chris Bervoets contributed equally to this work.

Conceptualization & Methodology: KL, CB, BN, LL; Analysis: KL, LL; Investigation: KL, CB, CD, LL; Data Curation: KL, CB, LL; Writing – Original Draft: KL, LL; Writing – Review & Editing: KL, CB, CD, BN, LL; Visualization: LL; Supervision: BN, LL; Project administration: KL; Funding acquisition: BN, LL. All authors read and approved the final manuscript.

## Acknowledgements

We are grateful for the patients’ time and effort to participate in this study. We thank Mathijs Franssen and Jeroen Clarysse for their help with programming the computerized symptom provocation task in Affect 5.0, John Das for his help with programming the patients’ stimulation parameters, Brecht Decraene and Asuka Nakajima for their input regarding the electrode contact selection, and Loes Gabriëls for her input throughout the study. We acknowledge the financial support of the Research Foundation – Flanders (FWO) (Research Project G0C9817N), Medtronic Chair for Stereotactic Neurosurgery in Psychiatric Disorders (B. Nuttin), European Research Council (CoG 648176), and KU Leuven Research Council (Project 3H190245).

## SUPPLEMENT

### Participant flow

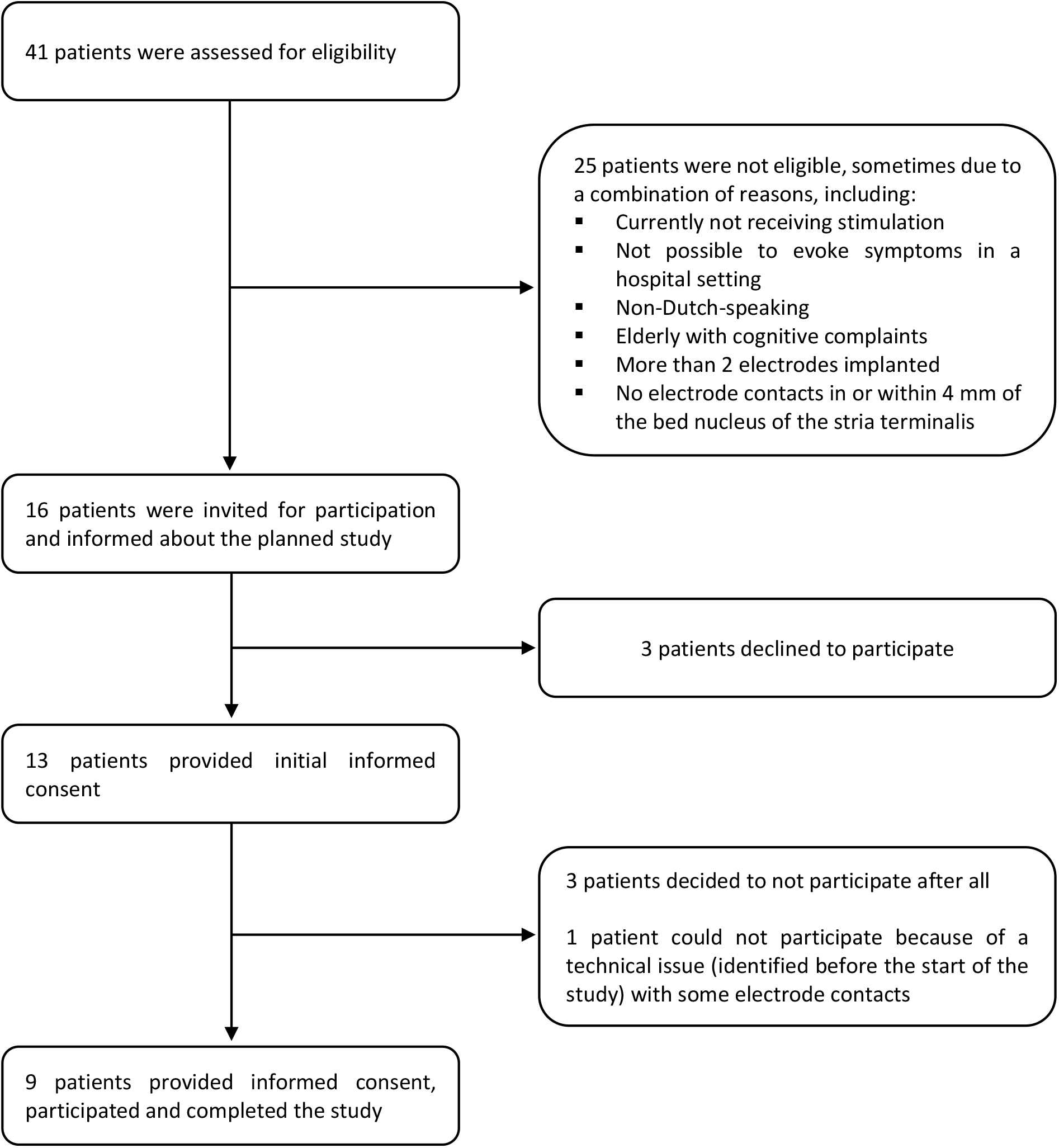

### Sample characteristics

**Table S1:** Pre-implantation measurements.

| PATIENT | OCD SUBTYPE | Y-BOCS Preop |
| --- | --- | --- |
| Pt01 | CHECK | 34 |
| Pt02 | WASH | 36 |
| Pt03 | WASH | 40 |
| Pt04 | CHECK | 32 |
| Pt05 | CHECK | 35 |
| Pt06 | WASH | 36 |
| Pt07 | WASH | 39 |
| Pt08 | CHECK | 32 |
| Pt09 | WASH | 36 |

**Table S2:**
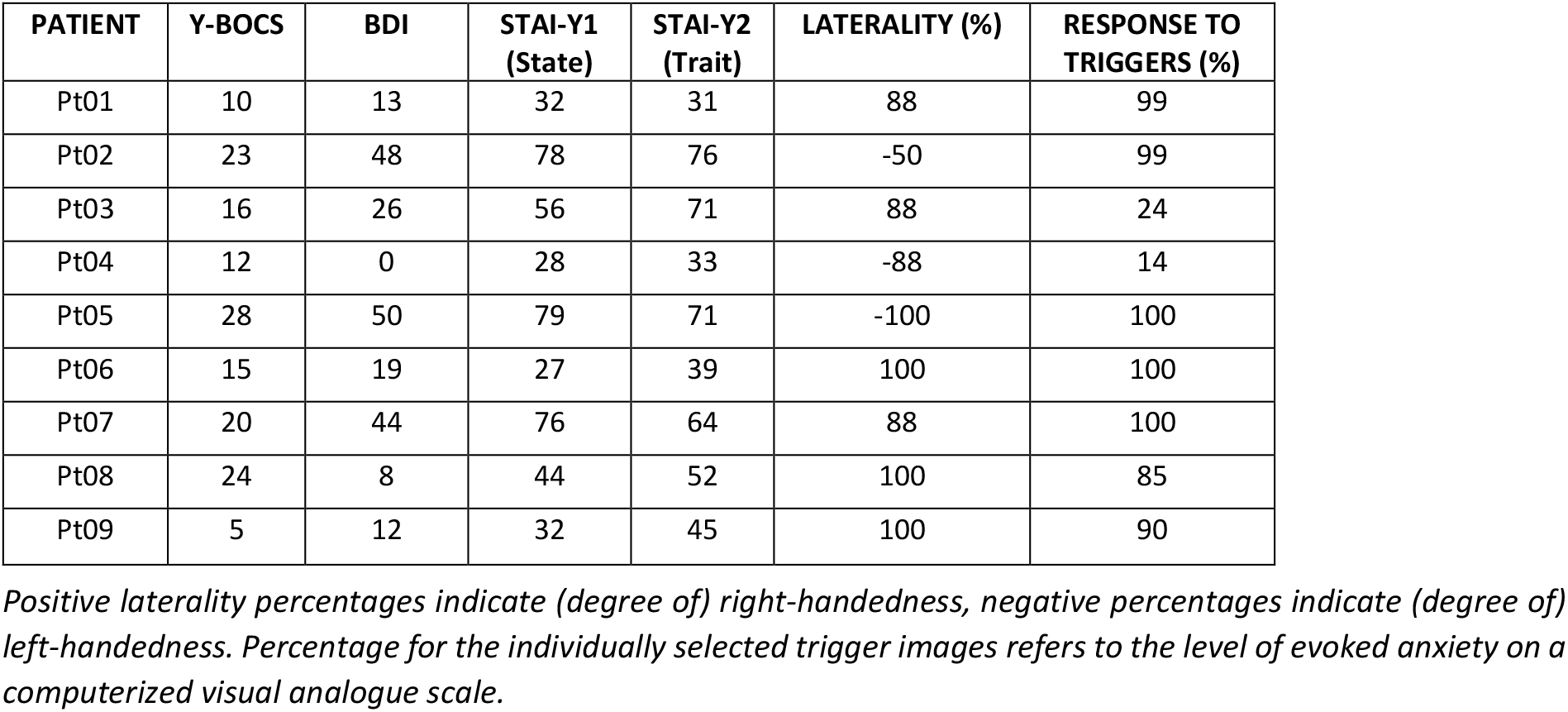
Baseline measurements (collected maximally 10 weeks before symptom provocation)

**Fig. S1:**
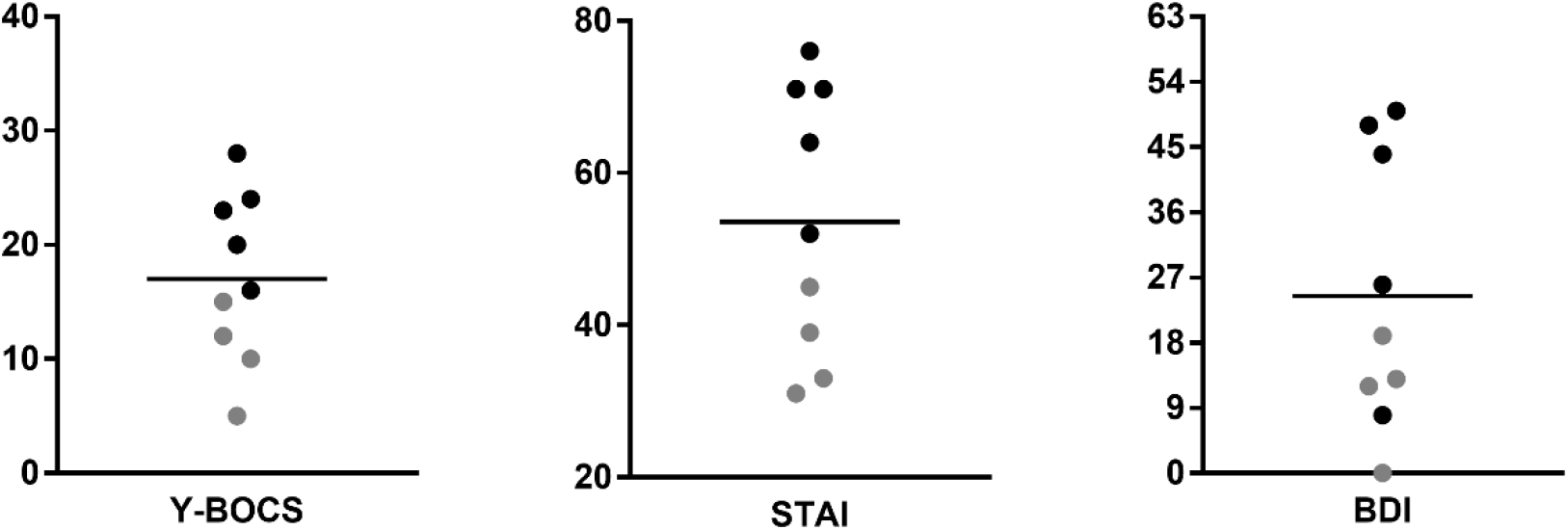
Baseline measurements of obsessions and compulsions (Y-BOCS), anxiety (STAI-Trait) and depression (BDI) (collected maximally 10 weeks before the symptom provocation task) Individual data (and means) of 9 patients are shown for measurements collected before the start of the symptom provocation task. Patients with mild OCD (Y-BOCS ≤ 15) are indicated with gray symbols, patients with moderate-severe OCD (Y-BOCS > 15) are depicted with black symbols. Y-BOCS: Yale-Brown Obsessive Compulsive Scale; STAI: State-Trait Anxiety Inventory; BDI: Beck Depression Inventory.

### Stimulation characteristics

**Table S3:** Contacts used for BST stimulation & regular stimulation parameters.

|  |  |  | BST STIMULATION CONTACTS |  | REGULAR STIMULATION PARAMETERS<br>(APPLIED DURING HAB PHASE) |  |  |  |  | THERAPEUTIC IMPEDANCE |  |
| --- | --- | --- | --- | --- | --- | --- | --- | --- | --- | --- | --- |
| PATIENT | LEAD TYPE | SIDE | CATHODE (-) | ANODE (+) | CATHODE (-) | ANODE (+) | FREQUENCY (Hz) | PULSE WIDTH (μs) | VOLTAGE (V) | (Ohm) | (mA) |
| Pt01 | 3387 | LEFT | C1 | C2 | C0; C1 | case | 130 | 210 | 3 | 557 | 5.3 |
|  |  | RIGHT | C1 | C2 | C0; C1 | case | 130 | 210 | 3 | 673 | 4.4 |
| Pt02 | 3391 | LEFT | C0 | C1 | C0; C1 | C2 | 130 | 240 | 4.1 | N/A | N/A |
|  |  | RIGHT | C0 | C1 | C0; C1 | C2 | 130 | 240 | 5.1 | N/A | N/A |
| Pt03 | 3391 | LEFT | C0 | C1 | C1; C2 | case | 130 | 240 | 4.6 | 428 | 10.3 |
|  |  | RIGHT | C0 | C1 | C0; C1 | case | 130 | 240 | 4.6 | 462 | 9.5 |
| Pt04 | 3391 | LEFT | C0 | C1 | C1 | case | 130 | 210 | 4.5 | 506 | 8.6 |
|  |  | RIGHT | C0 | C1 | C1 | case | 130 | 210 | 4.5 | 545 | 8.1 |
| Pt05 | 3391 | LEFT | C0 | C1 | C0; C1 | case | 190 | 400 | 1.6 | 597 | 2.5 |
|  |  | RIGHT | C0 | C1 | C0; C1 | case | 190 | 400 | 2 | 563 | 3.3 |
| Pt06 | 3391 | LEFT | C0 | C1 | C1 | case | 130 | 180 | 7 | 769 | 9 |
|  |  | RIGHT | C0 | C1 | C1 | case | 130 | 180 | 7 | 806 | 8.6 |
| Pt07 | 3391 | LEFT | C0 | C1 | C0 | C3 | 170 | 200 | 4 | 1023 | 3.9 |
|  |  | RIGHT | C0 | C1 | C0 | C3 | 170 | 200 | 4 | 1275 | 3.2 |
| Pt08 | 3391 | LEFT | C0 | C1 | C0; C1 | C3 | 170 | 300 | 5.5 | 589 | 8.8 |
|  |  | RIGHT | C0 | C1 | C0; C1 | C3 | 170 | 300 | 5.5 | 583 | 9 |
| Pt09 | 3391 | LEFT | C0 | C1 | C1 | case | 130 | 240 | 3 | 210 | 5.8 |
|  |  | RIGHT | C0 | C1 | C1 | case | 130 | 240 | 3 | 582 | 5.1 |

**Table S4.**
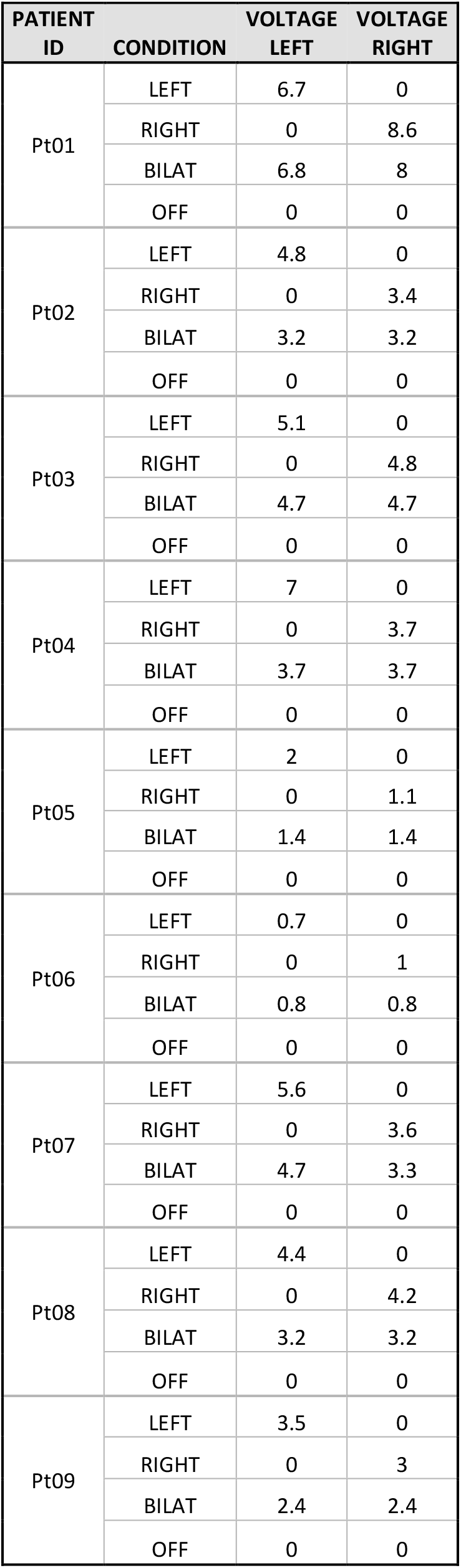
Stimulation voltages during crossover phase.

| PATIENT ID | CONDITION | VOLTAGE LEFT | VOLTAGE RIGHT |
| --- | --- | --- | --- |
| Pt01 | LEFT | 6.7 | 0 |
|  | RIGHT | 0 | 8.6 |
|  | BILAT | 6.8 | 8 |
|  | OFF | 0 | 0 |
| Pt02 | LEFT | 4.8 | 0 |
|  | RIGHT | 0 | 3.4 |
|  | BILAT | 3.2 | 3.2 |
|  | OFF | 0 | 0 |
| Pt03 | LEFT | 5.1 | 0 |
|  | RIGHT | 0 | 4.8 |
|  | BILAT | 4.7 | 4.7 |
|  | OFF | 0 | 0 |
| Pt04 | LEFT | 7 | 0 |
|  | RIGHT | 0 | 3.7 |
|  | BILAT | 3.7 | 3.7 |
|  | OFF | 0 | 0 |
| Pt05 | LEFT | 2 | 0 |
|  | RIGHT | 0 | 1.1 |
|  | BILAT | 1.4 | 1.4 |
|  | OFF | 0 | 0 |
| Pt06 | LEFT | 0.7 | 0 |
|  | RIGHT | 0 | 1 |
|  | BILAT | 0.8 | 0.8 |
|  | OFF | 0 | 0 |
| Pt07 | LEFT | 5.6 | 0 |
|  | RIGHT | 0 | 3.6 |
|  | BILAT | 4.7 | 3.3 |
|  | OFF | 0 | 0 |
| Pt08 | LEFT | 4.4 | 0 |
|  | RIGHT | 0 | 4.2 |
|  | BILAT | 3.2 | 3.2 |
|  | OFF | 0 | 0 |
| Pt09 | LEFT | 3.5 | 0 |
|  | RIGHT | 0 | 3 |
|  | BILAT | 2.4 | 2.4 |
|  | OFF | 0 | 0 |

**Fig. S2.**
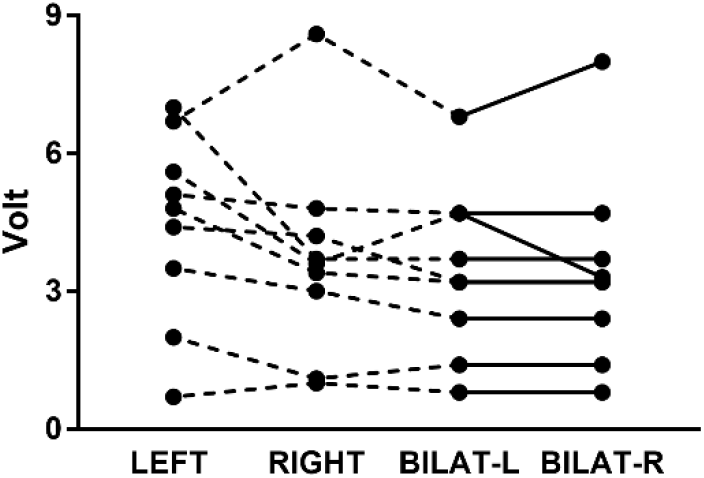
The figure above shows individual voltages of 9 patients for electrical stimulation of the BST with a fixed frequency of 130 Hz and pulse width of 450 µs. Bilateral stimulation voltages in the left (BILAT-L) and right (BILAT-R) hemisphere are connected with a solid line. The highest tolerable amplitude (HTA) for stimulation of the left (LEFT), right (RIGHT) and bilateral BST was determined before the start of the crossover phase. Left and right HTAs were determined in random order, always followed by selection of the bilateral HTA. As mentioned in the main text, the stimulation amplitude was gradually increased to determine the HTA. This was typically done using increments of 0.2-0.5 V in 30-s steps. When non-transient side effects were observed, the amplitude was reduced to ±90% and evaluated for tolerance. A one-way repeated-measures ANOVA with Greenhouse-Geisser correction was conducted with pre-specified comparisons (Holm-Sidak’s multiple comparions tests). The ANOVA showed a trend toward a main effect of hemisphere (F(1.41,11.28) = 3.84, p = .07). We found no statistically significant differences between stimulation voltages applied in the left versus right hemisphere (p = .18). We found trends toward lower voltages with bilateral versus unilateral stimulation (left hemisphere: p = .06, right hemisphere: p = .09).

**Table S5:** Questions used for VAS scales (Dutch & English translation)

| **Original (Dutch) version** |
| --- |
| **Introductory sentences**<br>Before neutral images: “U gaat nu enkele foto’s te zien krijgen.”<br>Before trigger images for checkers: “Beeld u in dat u de volgende situaties niet kan controleren.”<br>Before trigger images for washers: “Beeld u in dat u de volgende zaken niet kan proper maken.” |
| **Next, a sequence of 5 personalized trigger images is presented, followed by 6 VAS scales:**<br>Q1: In hoeverre voel je je nu angstig/gespannen? (Helemaal niet ---> Zeer uitgesproken)<br>Q2: In hoeverre voel je je nu neerslachtig? (Helemaal niet ---> Zeer uitgesproken)<br>Q3: In welke mate heeft u nu last van dwanggedachten? (Helemaal niet ---> Zeer uitgesproken)<br>Q4: Hoe sterk voel je nu de neiging tot het stellen van een dwanghandeling? (Helemaal niet ---> Zeer uitgesproken)<br>Q5: In hoeverre voel je de neiging deze situatie te vermijden? (Helemaal niet ---> Zeer uitgesproken)<br>Q6: Wat is momenteel je algemeen gevoel van welbevinden? (Zeer slecht ---> Zeer goed) |
| **English translation** |
| **Introductory sentences**<br>Before neutral images: “You will now see a few pictures.”<br>Before trigger images for checkers: “Imagine that you cannot check the following situations.”<br>Before trigger images for washers: “Imagine that you cannot clean the following items.” |
| **Next, a sequence of 5 personalized trigger images is presented, followed by 6 VAS scales:**<br>Q1: To what extent are you now feeling anxious/tense? (Not at all ---> Very much)<br>Q2: To what extent are you now feeling depressed? (Not at all ---> Very much)<br>Q3: To what extent are you now experiencing obsessive thoughts? (Not at all ---> Very much)<br>Q4: How strong are you now feeling the urge to perform compulsions? (Not at all ---> Very much)<br>Q5: To what extent do you feel the urge to avoid this situation? (Not at all ---> Very much)<br>Q6: What is your overall feeling of wellbeing at this moment? (Very bad ---> Very good) |

**Fig. S3:**
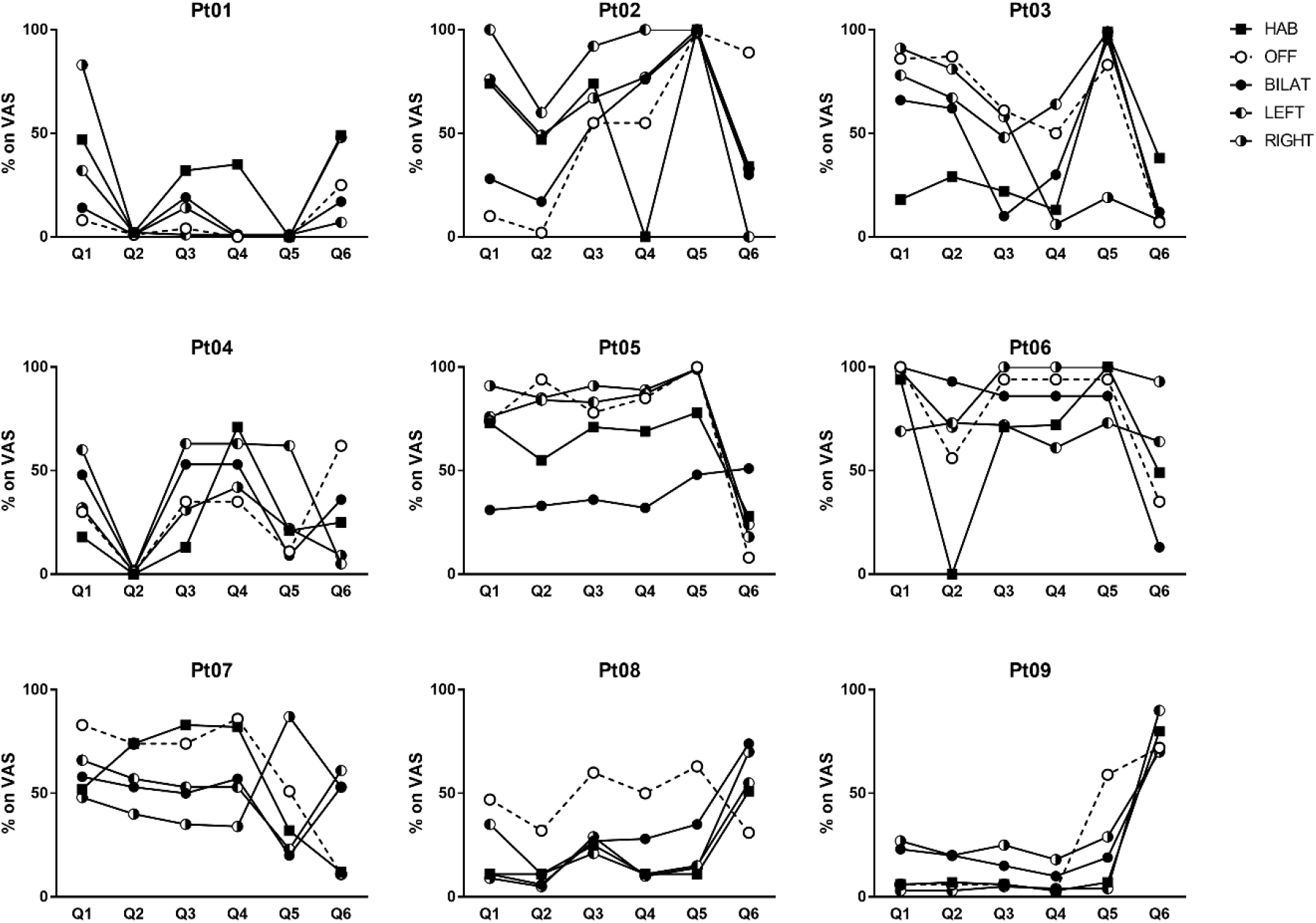
Responses on VAS scales - Individual patient data. Individual data of 9 patients are shown for the responses to 6 questions related to feelings of anxiety/stress (Q1), depression (Q2), obsessions (Q3), compulsions (Q4), tendency to avoid (Q5) and feeling of wellbeing (Q6). Patients first underwent an open-label Habituation (HAB) phase during which they received stimulation as usual. During the crossover, patients went through 4 double-blind, randomized phases (OFF (no stimulation), BILAT (bilateral), LEFT and RIGHT stimulation). Symbols for the OFF condition are shown in white since it is the only condition without any electrical brain stimulation. VAS: visual analogue scale.

**Fig. S4:**
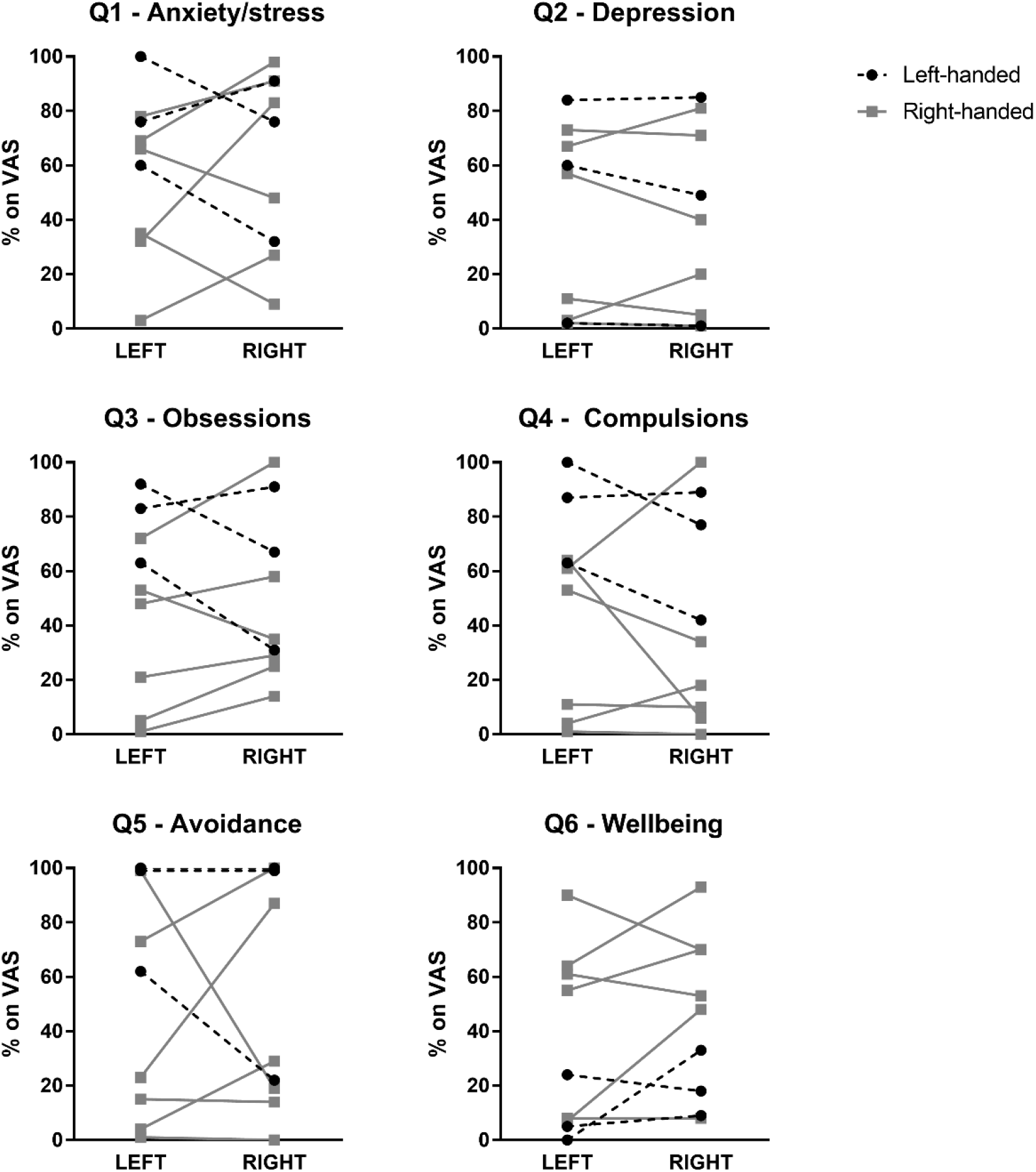
Laterality. For exploratory purposes, we compared responses on the VAS scales with left BST stimulation versus right BST stimulation for left-handed (n = 3) versus right-handed (n = 6) patients. We conducted two-way repeated-measures ANOVAs for each question and were mainly interested in interactions between handedness and stimulated hemisphere, none of which were statistically significant (F(1,7) = 1.55, p = .25 for Q1, F(1,7) = 0.32, p = .59 for Q2, F(1,7) = 4.61, p = .07 for Q3, F(1,7) = 0.23, p = .65 for Q4, F(1,7) = 0.40, p = .55 for Q5 and F(1,7) = 0.003, p = .96 for Q6).

### Electrocardiography (ECG)

ECG electrodes (Kendall H66LG, 55 mm diameter) were attached to the right and left mid-clavicle ((-) and ground, respectively) and lower left rib cage (+) of the patient. The ECG signal was recorded at 1 kHz and low-pass filtered below 150 Hz (LabLinc V model V75-04, Coulbourn Instruments, Allentown, Pennssylvania). Digital conversion was done using a National Instruments data acquisition system (16-bit PCI-6221 card with NI BNC-2111 connector block, National Instruments, Austin, Texas) and Affect 5.0. Peaks were automatically detected using a custom-made Matlab algorithm. Quality of peak detection was manually evaluated for every segment. In case of insufficient quality, peaks were manually detected by a blinded observer using a custom-made Matlab GUI.

Data from one patient (Pt08) were lost due to technical difficulties. We extracted 45-s segments of the ECG signal that coincided with presentation of each series of neutral and trigger images, during all five stimulation conditions (HAB, OFF, BILAT, LEFT, RIGHT). Comparison of ECG measurements during neutral versus trigger images is limited by the fact the order was not counterbalanced; neutral was always shown first. For every image type (neutral or trigger), we calculated weighted RMSSD (Root Mean Square of Successive Differences) and inter-beat intervals (then converted to heart rate in beats per minute (bpm).

**Fig. S5:**
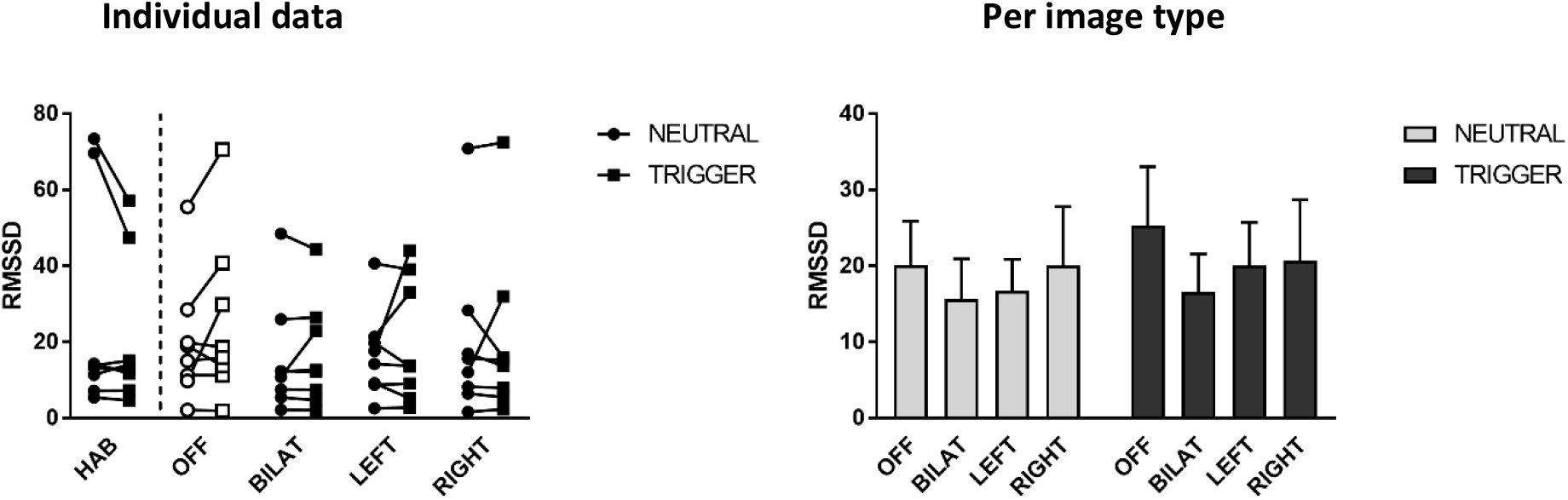
Heart rate variability (RMSSD) To assess whether the two image types (neutral and trigger) had an effect on heart rate variability while the patient was being stimulated with their regular parameters (HAB phase), we conducted a paired t-test and found no significant difference (t(7) = 1.47, p = .19). To evaluate differences during the crossover trial (comparing OFF, BILAT, LEFT and RIGHT; right panel of the figure shows means and SEM), we conducted a two-way repeated-measures ANOVA, and found no main effects of image type (F(1,7) = 1.36, p = .28), or stimulation condition (F(3,21) = 0.59, p = .63), nor interaction (F(3,21) = 0.71, p = .55). In conclusion, we found no significant differences in heart rate variability using 45-s measurements during neutral and trigger images in the different stimulation conditions.

**Fig. S6:**
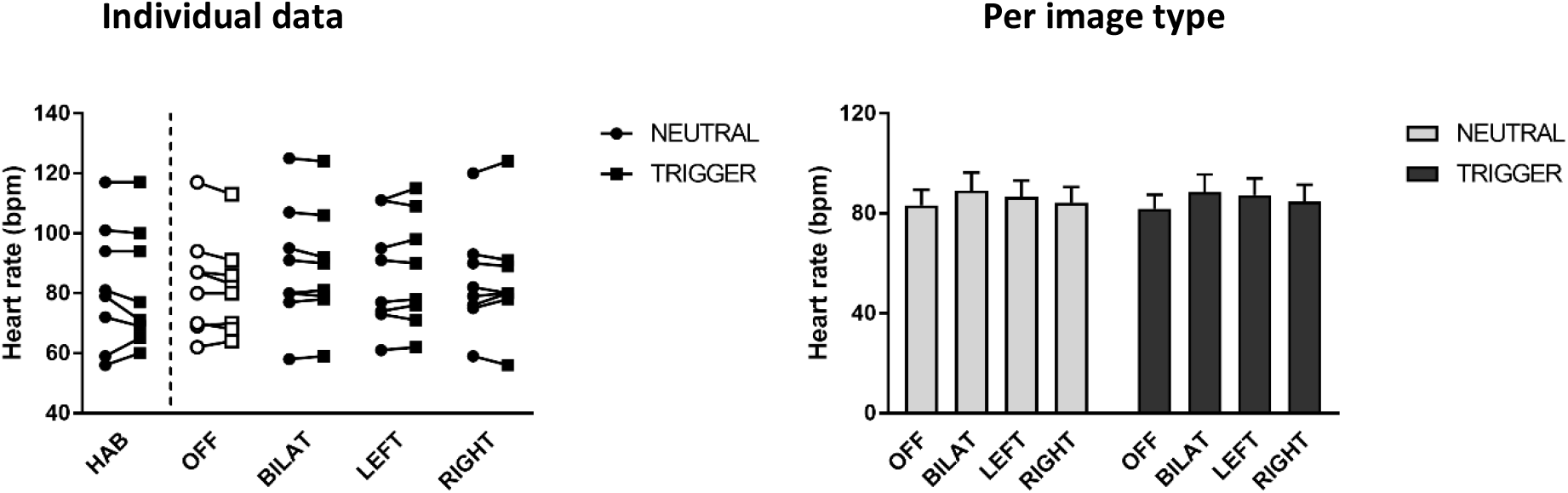
Heart rate (bpm) To assess whether the two image types (neutral and trigger) had an effect on heart rate while the patient was being stimulated with their regular parameters (HAB phase), we conducted a paired t-test and found no significant difference (t(7) = 0.48, p = .65). To evaluate differences during the crossover trial (comparing OFF, BILAT, LEFT and RIGHT; right panel of the figure shows means and SEM), we conducted a two-way repeated-measures ANOVA, and found no main effects of image type (F(1,7) = 0.12, p = .73), or stimulation condition (F(3,21) = 1.36, p = .28), nor interaction (F(3,21) = 1.61, p = .22). In conclusion, we found no significant differences in heart rate using 45-s measurements during neutral and trigger images in the different stimulation conditions.

